# Perioperative Patient Blood Management (PBM) in Africa: a multicentre mixed-methods assessment of health system readiness and clinical practice in Ethiopia

**DOI:** 10.64898/2026.07.30.26359311

**Authors:** Fitsum K Belachew, Tesfay Yohannes Ambese, Peniel Kenna Dulla, Tariku Assefa, Abiy Sisay, Abiy Dawit, Fredy Ariza, Jolene Moore

## Abstract

**Background:** Perioperative Patient Blood Management (PBM), a patient-centred, evidence-based approach to improve outcomes by preserving and optimising a patient’s own blood, remains unimplemented in most low- and middle-income countries. Few studies have assessed health-system readiness for PBM implementation, anaemia management, or transfusion practices in sub-Saharan Africa. This study provides a pre-implementation baseline assessment of perioperative PBM readiness in Ethiopia.

**Materials and Method:** A mixed-methods approach evaluated 12 public hospitals in Addis Ababa, with the PBM Facility Assessment Tool to determine readiness scores. Data from 1,236 surgical patients assessed anaemia, transfusions, and predictors. Interviews with clinicians, blood bank staff, and policymakers explored decision-making and barriers. Findings were integrated through triangulation.

**Results:** The composite PBM-FAT score was 6.6 ± 0.7 (range 5.5–7.5); however, this reflects general perioperative and transfusion-service infrastructure rather than PBM-specific capability. Domain analysis revealed critical gaps: 75% of facilities lacked intravenous iron, viscoelastic testing was absent in all, and 66.7% lacked point-of-care haemoglobin testing. Fibrinogen and haematinic tests were rarely available (66–92% never tested). Preoperative anaemia was documented in 7.1% of patients, a minimum estimate reflecting detection failure due to non-systematic haemoglobin testing, and perioperative transfusion occurred in 2.4%, more consistent with supply rationing than with optimised clinical practice. Independent transfusion predictors included cancer surgery (AOR 11.28, 95% CI 3.00–42.36) and blood loss ≥500 mL (AOR 9.76, 95% CI 3.94–24.17). Interviews revealed blood shortages, the absence of national PBM governance, transfusion as the default treatment for anaemia, and highly variable thresholds.

**Conclusion:** Ethiopian hospitals have functional transfusion services but lack the anaemia management pathways, diagnostics, therapeutics, and governance structures required for Patient Blood Management. PBM is not yet operationalised as a patient-centred, preventive care strategy. Priority implementation steps, aligned with the WHO 2024 PBM guidance, include systematic preoperative haemoglobin screening, access to intravenous iron, blood utilisation governance, and a national PBM framework to improve patient outcomes.

**Visual summary:** 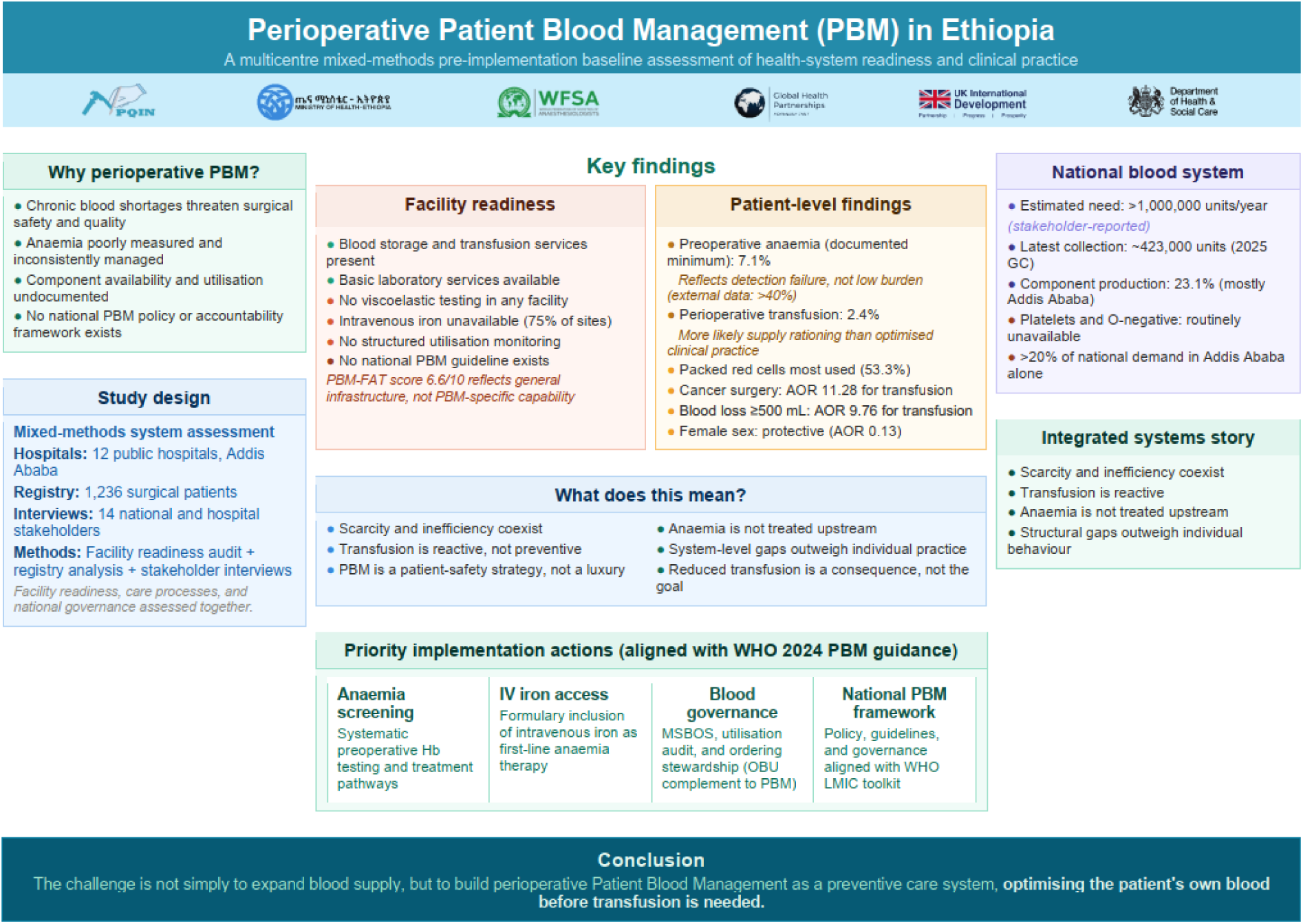

**Highlights:**

- Ethiopian public hospitals have functional transfusion and perioperative infrastructure (composite PBM-FAT score 6.6/10) but lack the PBM-specific capabilities, systematic anaemia management, intravenous iron, and haemostatic diagnostics required for patient-centred blood optimisation.
- Systemic barriers, including blood shortages, the absence of national PBM governance, and inconsistent practices, highlight the urgent need for national guidelines and utilisation monitoring to strengthen surgical safety.
- Blood shortages coexist with inefficient utilisation practices, indicating that governance gaps compound supply constraints.

## Introduction

Patient blood management (PBM) is a patient-centred, systematic, evidence-based approach to improve patient outcomes by managing and preserving a patient’s own blood, while promoting patient safety and empowerment (1). Reduced exposure to allogeneic blood transfusion is an important downstream consequence of PBM rather than its primary goal. PBM is built around three core pillars: optimisation of the patient’s own blood (anaemia and iron deficiency management), minimisation of perioperative blood loss, and rational use of allogeneic transfusion only when clinically indicated (2). In high-income countries, PBM programs have been widely implemented and shown to reduce transfusion exposure, morbidity, mortality, length of hospital stays, and healthcare costs (3).

However, PBM adoption in low- and middle-income countries (LMICs) remains limited. The Lancet Commission on Global Surgery highlighted major inequalities in surgical safety and outcomes globally, with more than 4.2 million people dying within 30 days of surgery each year, most in LMICs (4). A systematic review similarly reported that although PBM is recognised as an effective intervention, implementation is often constrained by inadequate infrastructure, limited clinician awareness, and shortages of essential resources (3). These challenges are particularly acute in sub-Saharan Africa, where anaemia is highly prevalent, and national blood supplies frequently fall short of clinical demand (5,6).

In Ethiopia, the burden of surgically treatable diseases is high, and postoperative complications contribute substantially to avoidable morbidity and mortality (7,8). The Ethiopian Surgical Outcomes Study (Ethio-SOS) found that nearly one in five surgical patients experienced a postoperative complication, with bleeding and reoperation among the leading contributors (8). Yet empirical data on the prevalence of preoperative anaemia, perioperative transfusion practices, and facility readiness for PBM remain scarce. A recent national Service Availability and Readiness Assessment (SARA) of Ethiopian health facilities reported that fewer than 5% met the criteria for full transfusion service readiness, highlighting major gaps in laboratory capacity, blood product availability, and standardised protocols (9). These systemic challenges highlight the urgent need for locally grounded evidence to inform PBM implementation strategies.

Accordingly, this study was designed to provide a comprehensive pre-implementation baseline assessment of health-system readiness for perioperative Patient Blood Management (PBM) in Ethiopia using a mixed-methods approach. First, we evaluated facility-level readiness for PBM, including laboratory capacity, blood product availability, workforce, equipment, and the presence of relevant policies and protocols. Second, we quantified the burden of preoperative anaemia and characterised perioperative transfusion patterns using multicentre perioperative registry data. Third, we explored provider decision-making, transfusion thresholds, and perceived barriers to PBM through interviews with anaesthesia care providers, surgeons, and blood bank personnel. Finally, we integrated these datasets to identify overarching gaps and opportunities to inform the phased nationwide implementation of PBM.

## Materials and Methods

### Study design

The study used a mixed-methods design to provide a pre-implementation baseline assessment of health-system readiness for perioperative Patient Blood Management (PBM) in Addis Ababa, Ethiopia’s hub for national referral hospitals, blood services, and surgical volume. The quantitative component assessed facility readiness using the standardised PBM Facility Assessment Tool (PBM-FAT), which evaluated laboratory capacity, blood products, policies, workforce, medications, equipment, and perioperative services. The PBM-FAT used in this study was developed by the World Federation of Societies of Anaesthesiologists (WFSA) as a pilot instrument for perioperative PBM readiness. Its structure was derived from the WHO Patient Blood Management guideline(10), operationalising WHO recommendations into a practical facility-level checklist suitable for LMIC surgical systems. Although WHO does not publish a standalone PBM readiness tool, the PBM-FAT aligns closely with the WHO PBM pillars and system-level requirements. The full PBM-FAT is provided in Suppementary material 2.

This was complemented by secondary analysis of perioperative registry data to examine preoperative anaemia patterns, transfusion practices, and outcomes. Semi-structured qualitative interviews explored decision-making, thresholds, blood conservation, and barriers to PBM among anaesthesia providers, surgeons, and blood bank staff. The observational component of the study is reported in accordance with the Strengthening the Reporting of Observational Studies in Epidemiology guideline (STROBE)(11); the qualitative component was reported with reference to the Consolidated Criteria for Reporting Qualitative Research (COREQ)(12), and mixed-methods integration is reported with reference to the Good Reporting of A Mixed Methods Study (GRAMMS) framework(13).

### Study population

The study population comprised three groups:

- Facility readiness arm: hospitals in Addis Ababa, including secondary and tertiary referral centres that provide care to patients from across the country and operate under national blood supply and policy frameworks.
- Quantitative arm: all surgical patients recorded in the Perioperative Registry between 1 January and 30 September 2025 from Addis Ababa hospitals participating in the national perioperative registry network.
- Qualitative arm: a purposive sample of anaesthesia providers, surgeons, blood bank personnel, and national stakeholders involved in perioperative or transfusion-related decision-making, including hospital leaders, the Ministry of Health and Ethiopian blood and tissue bank leaders.

Inclusion and exclusion criteria

Inclusion criteria were surgical patients with complete data on preoperative haemoglobin, transfusion status, and discharge outcome; for the qualitative study, clinicians involved in perioperative or transfusion care. Exclusion criteria included incomplete surgical records, non-clinical status, or refusal to participate in the interview.

### Data sources

Data were obtained from three sources: first, PBM-FAT evaluations from the Ethiopian Surgical Outcome Study (Ethio-SOS), with Addis Ababa follow-ups via on-site visits/phone verification; second, perioperative registry (Version 2.4) data on demographics, ASA status, preoperative haemoglobin, blood loss, transfusions (type/units), complications, ICU admission, length of stay, and mortality; and third semi-structured interviews focusing on anaemia screening, transfusion thresholds, conservation strategies, decision-making, and PBM barriers. National DHIS2 data on blood related surgical cancellations were independently extracted and analysed by the study team.

### Outcomes

The primary outcomes were:

- Prevalence of preoperative anaemia
- Rate of perioperative blood transfusion Secondary outcomes included:
- Postoperative complications
- ICU admission
- In-hospital mortality
- Facility-level PBM readiness
- Qualitative themes Sample size calculation

The patient-level sample size was calculated using a single-proportion formula at 95% confidence. An expected perioperative transfusion prevalence of 22.7% was assumed based on previous perioperative studies (14) , with a ±3% margin of error, 1.5 design effect for hospital clustering, and 10% adjustment for missing data, yielding an estimated sample size of 1,236 patients. This sample size was selected to improve precision and support multivariable logistic regression analyses.

### Sampling procedure

Registry data (January-September 2025) covered 8,174 patients from five Addis Ababa hospitals. We selected 1,236 using proportionate stratified random sampling by hospital caseload (**Table 1**; **Figure 1**). For qualitative, purposive sampling, 14 diverse stakeholders (4 surgeons, 3 anaesthesia providers, 2 National Blood Bank representatives, 1 hospital blood bank delegate, 2 leaders, 2 Ministry representatives) with ≥2 years’ experience were recruited. Data saturation was reached, as assessed by code repetition. The qualitative interviews and facility assessments were conducted across overlapping tertiary and referral hospital settings in Addis Ababa, while the registry analysis was based on data from five participating hospitals within the same broader health-system context.

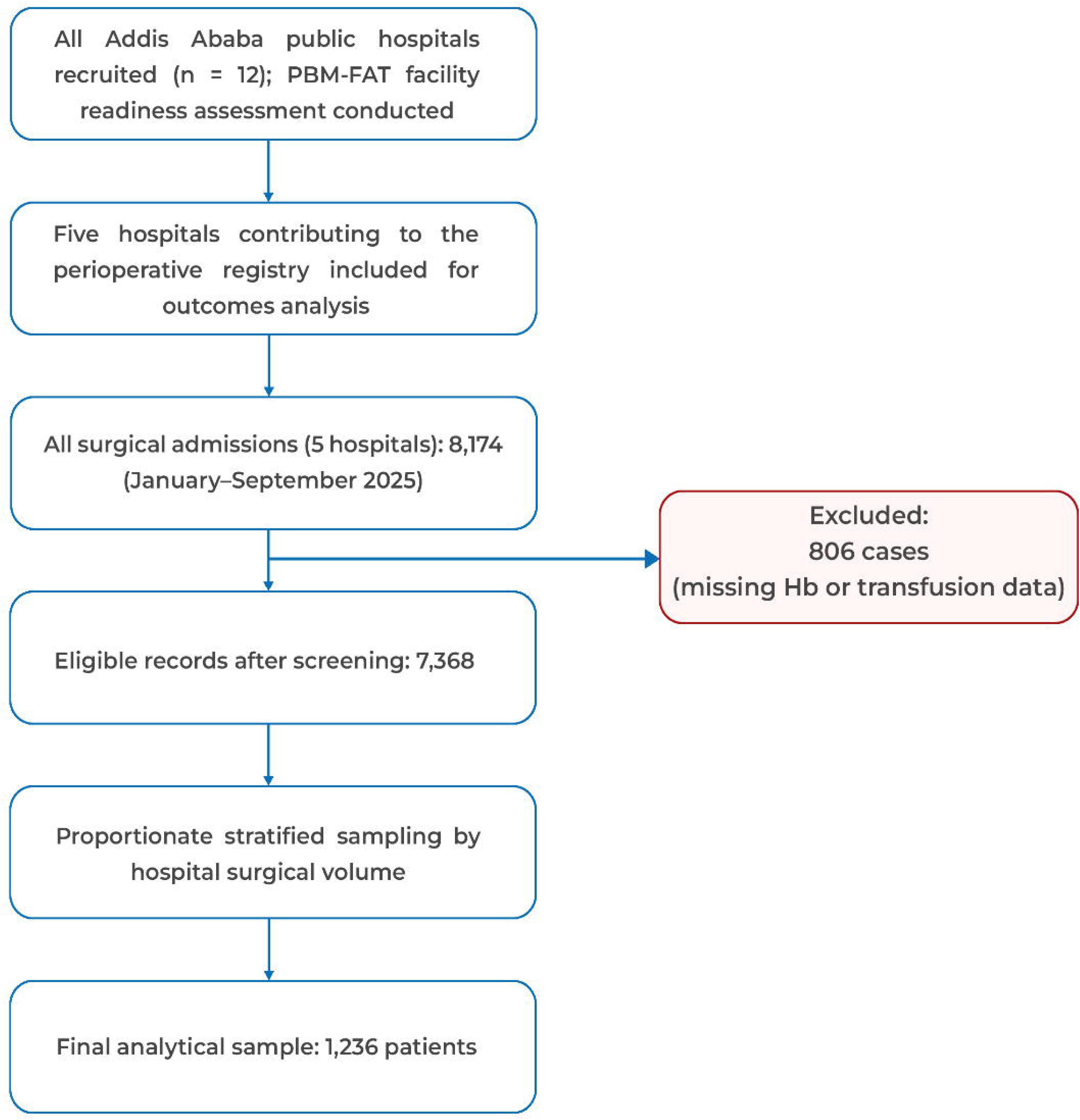

**Table 1:** Total surgical admissions and proportionate stratified sample sizes across participating hospitals (n = 1,236)

| Hospital code | Total surgical admissions | Sample size (n) |
| --- | --- | --- |
| Hospital 1 | 687 | 115 |
| Hospital 2 | 2050 | 344 |
| Hospital 3 | 656 | 110 |
| Hospital 4 | 1823 | 306 |
| Hospital 5 | 2152 | 361 |
| Total analytical sample | ---- | 1236 |

## Data analysis

### Quantitative analysis

Patient data from REDCap were analysed in Stata 17: categorical data as frequencies and percentages; continuous data for normality using the Kolmogorov-Smirnov test; comparisons using chi-square and T-tests. Multivariable logistic regression identified transfusion predictors, with adjusted odds ratios (AORs), 95% CIs, Hosmer-Lemeshow fit, and ROC discrimination. Anaemia was classified as per WHO criteria (see Supplementary Material **Table S2**)(15).

Facility readiness used PBM-FAT, administered April-June 2025, to key informants at 12 hospitals, verified against records. Each item was scored using a predefined algorithm implemented in Python: availability/frequency responses were assigned 3 (Always/Yes), 2 (More than half the time), 1 (Less than half the time), or 0 (Never/No/missing); time-based items (lab result turnaround, emergency RBC access) followed graduated scales (<30 min = 3, >4 hrs. = 0); and count-based items scored 3 if ≥1 and 0 otherwise. Raw item scores within each domain were summed and normalised to a 0–10 scale by dividing by the maximum possible domain score. An overall readiness score was computed as the unweighted mean of the nine domain scores, which were analysed, as detailed in **Table S1**. Descriptive statistics were used to summarise domain and overall scores across facilities. The facility assessment analysis was conducted using Python 3.13 with the Pandas and NumPy libraries.

### Qualitative analysis

Semi-structured interviews (n=14) were conducted in November 2025, audio-recorded, transcribed verbatim, and analysed using Braun and Clarke’s reflexive thematic analysis framework. Two members of the research team independently coded transcripts inductively in Taguette, followed by iterative comparison of codes, refinement of candidate themes, and resolution of discrepancies through discussion. Theme development followed the six phases of reflexive thematic analysis, moving from initial coding to theme generation, review, definition, and final reporting. To strengthen analytical validity, the team undertook reflexive discussions throughout coding and interpretation, drawing on their multidisciplinary backgrounds in perioperative quality improvement, surgical systems research, and transfusion-related implementation work in Ethiopia and other LMICs. These discussions were used to minimise individual bias and ensure consistency in theme development. Data saturation was assessed through code repetition, with no substantially new themes emerging in later interviews. No formal respondent validation (member checking) was undertaken. Representative quotations were incorporated to enhance transparency and credibility.

### Mixed-methods integration

Triangulation synthesised registry insights (anaemia and transfusions), PBM-FAT readiness, and interview barriers and facilitators into a narrative for policy implications.

### Ethical approvals

Ethical approval for this study was obtained from the Armauer Hansen Research Institute (AHRI) IRB (PO-64-23) for Ethio-SOS (including PBM-FAT) and Debre Berhan University Asrat Woldeyes Health Sciences Campus IRB (DBU-095) for registry data. Registry data were de-identified. Ministry support letters facilitated hospital participation. Qualitative participants provided audio-recorded verbal informed consent after a study explanation; data were stored securely in accordance with standards.

## Results

### Facility-level readiness for PBM

Twelve hospitals (6 secondary/general and 6 tertiary) in Addis Ababa were assessed using a structured PBM-FAT comprising nine domains, each scored from 0 to 10 (overall score = the mean of the domains). The overall readiness score was 6.6 ± 0.7 (range 5.5-7.5), as shown in **Table 2** and illustrated in **Figure 2**.

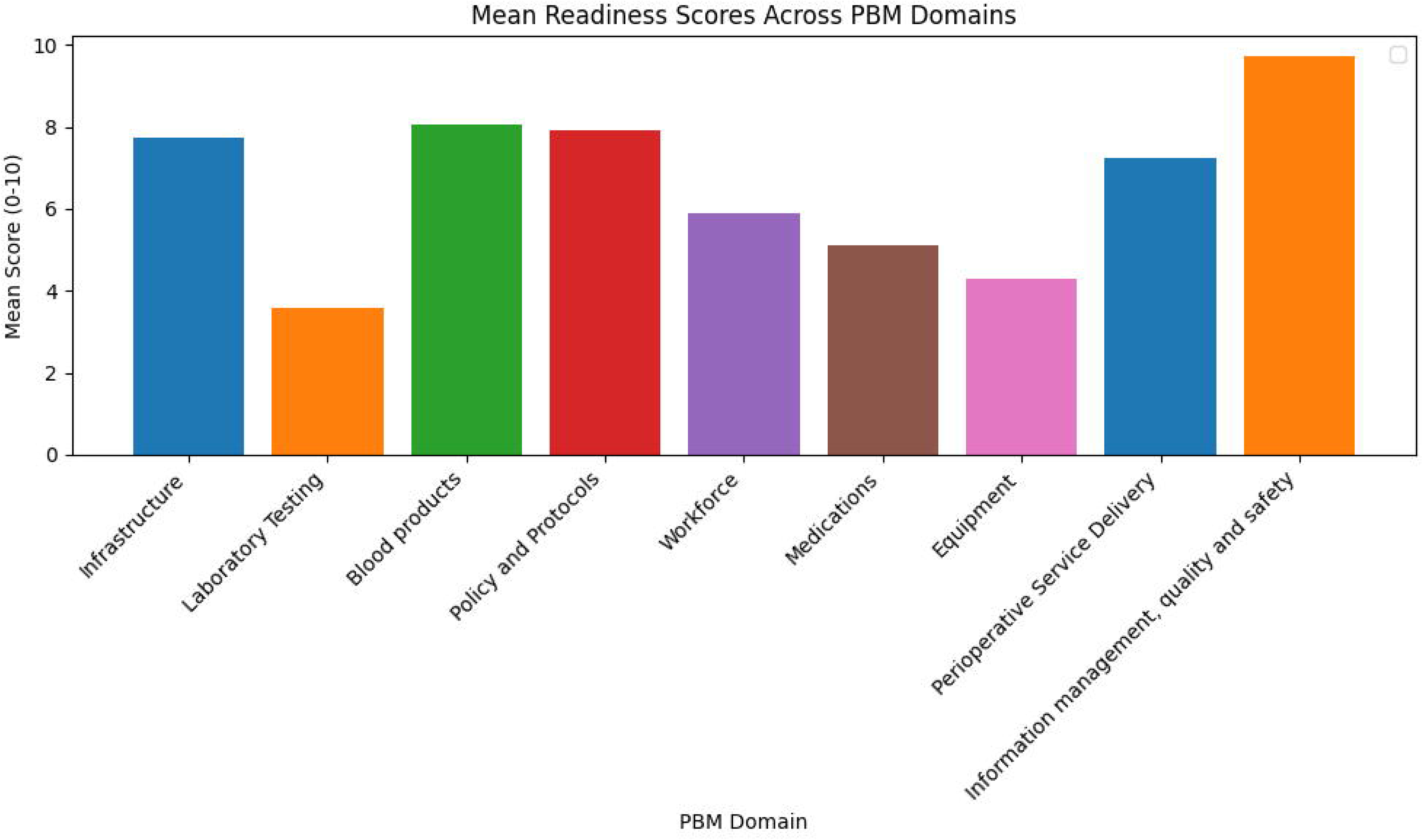

**Table 2:** Facility readiness scores for perioperative Patient Blood Management (PBM) implementation in Ethiopia.

| No | Domain | Mean score (SD) | Range |
| --- | --- | --- | --- |
| 1. | Infrastructure | 7.7 (0.8) | 6.7-8.6 |
| 2. | Laboratory testing | 3.6 (1.2) | 1.7-5.2 |
| 3. | Blood products | 8.1 (0.9) | 6.7-9.4 |
| 4. | Policy and protocols | 7.9 (2.4) | 3.3-10.0 |
| 5. | Workforce | 5.9 (1.0) | 3.9-7.2 |
| 6. | Medications | 5.1 (1.4) | 3.5-7.6 |
| 7. | Equipment | 4.3 (1.5) | 2.6-7.2 |
| 8. | Perioperative service delivery | 7.2 (0.5) | 6.2-7.9 |
| 9. | Information management, quality and safety | 9.7(0.7) | 8.3-10 |
|  | **Overall readiness score** | **6.6 (0.7)** | **5.5-7.5** |

Infrastructure had a mean score of 7.7 (SD 0.8), with 100% of facilities reporting oxygen availability and 91.7% reporting reliable electricity and backup power, blood products (8.1 ± 0.9); e.g., 100% had blood banks and typing; whole blood and packed RBCs always/mostly available in 83.3%, policy/protocols (7.9 ± 2.4), e.g., 100% monitored transfusion rates; 83.3% had hemovigilance committees, and perioperative services (7.2 ± 0.5); (e.g., 100% offered spinal/general anaesthesia and WHO Surgical Safety Checklist; major surgeries daily in 91.7%.

Laboratory testing had a mean score of 3.6 (SD 1.2). Fibrinogen level, haematinics (ferritin/folate/B12), serum iron, TIBC, and transferrin saturation are frequently not tested (66–92% never tested). In addition, viscoelastic testing was unavailable in all facilities, and point-of-care testing was unavailable in 66.7%), medications (5.1 ± 1.4) (e.g., basic vitamins and supplements such as oral folic acid (always 100%), IV vitamin B complex (always 100%), oral iron (always 83.3%), and IV/IM vitamin K (always 75– 83.3%) were widely available, while oral vitamin D (always only 41.7%), oral vitamin K (always only 8.3%), and erythropoiesis-stimulating agents (always only 25.0%) were much more limited or rarely present), and equipment (4.3 ± 1.5), (e.g., intra-arterial BP monitoring was never available in 66.7%; end-tidal CO2 was never available in 33.3%). Likewise, even simple items like warming devices and temperature measurement are not often available.

The workforce-specific staffing numbers include 408 surgeons, 677 midwives, 392 lab technicians, and specialised roles, such as transfusion medicine physicians, for a total of 4. Surgical volumes were 83,085 cases in the past 12 months, including 27,998 C-sections. Regarding information management, 91.7% had monthly QI meetings and morbidity/mortality reports. Detailed domain breakdowns are provided in **Table S3-S12.**

### Patient-level outcomes and transfusion practices

A total of 1,236 surgical patients were prospectively evaluated across the five hospitals. Mean age was 29.7 ± 15.4 years, 69.4% were female, and 48.6% underwent emergency procedures. The most common indications were non-communicable diseases (40.2%) and caesarean section (37.1%). Documented preoperative anaemia (WHO criteria) was identified in 88 patients (7.1%): mild 4.7%, moderate 2.1%, severe 0.3% (**Table 3**).

**Table 3:** Baseline characteristics of study participants.

| Characteristic | Overall<br>(N=1236) | With preoperative<br>anaemia (n=88) | Without<br>preoperative<br>anaemia (n= 1148) |
| --- | --- | --- | --- |
| Age in years, mean (SD) | 29.7 (15.4) | 33.1(17.0) | 29.4 (15.3) |
| Sex |  |  |  |
| Male | 378 (30.6%) | 37 (42.1%) | 341 (29.7%) |
| Female | 858 (69.4) | 51(57.9%) | 807 (70.3%) |
| ASA score, n (%) |  |  |  |
| I | 369 (29.8%) | 30(34.1%) | 339 (29.5%) |
| II | 745 (60.3%) | 44(50.0%) | 701(61.1%) |
| ≥ III | 122 (9.9%) | 14(15.9%) | 108 (9.4%) |
| Surgical Timing |  |  |  |
| Elective | 635 (51.4%) | 44 (50.0%) | 591 (51.5%) |
| Emergency | 601 (48.6%) | 44 (50.0%) | 557 (48.5%) |
| Indication of surgery |  |  |  |
| Noncommunicable<br>disease | 497 (40.2%) | 44(50.0%) | 453 (39.5%) |
| Caesarean section | 459 (37.1) | 18(20.5%) | 441 (38.4%) |
| Infection | 223 (18.0%) | 17(19.3%) | 206 (17.9%) |
| Trauma | 57(4.6%) | 9(10.2%) | 48 (4.2%) |
| Preoperative<br>haemoglobin, mean (SD), | 13.8(1.7) | 10.7 (1.8) | 14 (1.5) |
| g/dL |  |  |  |
| Cancer surgery |  |  |  |
| Yes | 33(2.7%) | 5 (5.7%) | 28 (2.4%) |
| No | 1203(97.3%) | 83(94.3%) | 1120(97.6%) |
| Anaemia category* |  |  |  |
| - Mild (Hb 10-12.9 g/dL) | 58 (4.7%) | 58 (4.7%) | N/A |
| - Moderate (Hb 7-9.9 g/dL) | 26(2.1%) | 26(2.1%) | N/A |
| - Severe (Hb <7 g/dL) | 4 (0.3%) | 4 (0.3%) | N/A |
| *Anaemia defined by WHO criteria; **Table S2**. SD: standard deviation; ASA: American Society of Anaesthesiologists; Hb: haemoglobin; NA: Not applicable. Data is presented in n (%). |  |  |  |

Any perioperative blood transfusion (regardless of the type of blood transfused) occurred in 29 patients (2.4%; 95% CI 1.6-3.4%). Intraoperative transfusion occurred in 1.2%, and postoperative transfusion in 1.5%. Packed red cells were used in 53.3% of transfusions, whole blood in 40.0%. Postoperative complications occurred in 1.5%, in-hospital mortality in 0.4%, and ICU admission in 0.4%. Median length of stay was 3 days (IQR 2-4) (**Table 4**).

**Table 4:** Perioperative blood transfusion and outcomes.

| Variables |  | Frequency (%) |
| --- | --- | --- |
| Transfusion | Intraoperative | 15 (1.2%) |
|  | Postoperative | 18 (1.5%) |
| Any transfusion * |  | 29(2.4%) |
| Type of blood transfused | Packed RBC | 8 (53.3%) |
|  | Whole blood | 6(40.0%) |
|  | Platelet | 1 (6.7%) |
| Post-op complication |  | 18 (1.5%) |
| In-hospital mortality |  | 5(0.4%) |
| ICU admission |  | 5 (0.4%) |
| Length of stay, median (IQR), days |  | 3(2-4) |
| *Any transfusion: either in intra-op or post-op transfused; RBC: Red blood cells, ICU: Intensive care unit; IQR: Interquartile range. |  |  |

In multivariable analysis, cancer surgery (AOR 11.28, 95% CI 3.00-42.36) and blood loss ≥500 mL (AOR 9.76, 95% CI 3.94-24.17) were independently associated with transfusion. Female sex was independently associated with a lower likelihood of transfusion (AOR 0.13, 95% CI 0.05–0.34). Preoperative anaemia showed a trend toward increased transfusion risk (AOR 2.64, 95% CI 0.93–7.52; p = 0.069). The model demonstrated good fit (Pearson χ²(584) = 476.95, Prob > χ² = 0.9996) and good discriminatory ability (area under the ROC curve = 0.8794) (**Table 5**).

**Table 5:** Univariable and multivariable logistic regression analyses for predictors of perioperative blood transfusion.

| Predictor |  | COR (95% CI) | AOR (95% CI) | P-value |
| --- | --- | --- | --- | --- |
| Preoperative anaemia | No | Ref |  |  |
|  | Yes | 3.58(1.42, 9.03) | 2.64 (0.93-7.52) | 0.069 |
| Age in years |  | 1.03(1.01, 1.05) | 1.01 (0.99-1.04) | 0.278 |
| Sex | Male | Ref |  |  |
|  | Female | 0.19(0.08, 0.42) | 0.13(0.05,0.34) | <0.001* |
| Surgery urgency | Emergency | Ref |  |  |
|  | Elective | 1.17(0.56,2.45) | 0.44 (0.18-1.09) | 0.076 |
| ASA status | ASA I | Ref |  |  |
|  | ASA II | 0.26 (0.12, 0.58) | 0.64(0.25, 1.66) | 0.361 |
|  | ASA ≥ III | 0.16 (0.02, 1.22) | 0.29(0.04,2.40) | 0.251 |
| Cancer surgery | No | Ref |  |  |
|  | Yes | 14.4 (5.67, 36.82) | 11.28(3.0, 42.36) | <0.001* |
| Estimated blood loss | < 500 mL | Ref |  |  |
|  | ≥ 500 mL | 7.16 (3.39, 15.15) | 9.76 (3.94-24.17) | <0.001* |
| COR: Crude odds ratio; AOR: Adjusted odds ratio; CI: Confidence interval. ASA: American society of anaesthesiologists; Ref: Reference; *Significant at p-value <0.05 |  |  |  |  |

### Qualitative findings

Fourteen in-depth interviews were conducted in November 2025 with key stakeholders: national blood bank leadership (n = 2), hospital blood bank (laboratory) (n = 1), anaesthesia providers (n = 3), surgeons (n = 4), hospital CEOs (n = 2), and ministry leaders (n = 2). Using Braun and Clarke’s thematic analysis framework, seven themes were identified.

***<u>National blood system capacity (reported by national blood service leadership):</u>*** respondents from the national blood service provided institutional estimates indicating that annual blood collection was approximately 423,000 units, whereas the national requirement exceeded 1 million units per year. Planned collection for the Ethiopian calendar (EC) 2018 (2025/2026 Gregorian Calendar) was reported as ≥550,000 units. Component preparation was reported to involve approximately 86,000 units (23.1%) annually, with processing limited primarily to Addis Ababa and typically dependent on case-specific requests.

Platelet availability was reported as critically constrained, with estimated demand at Tikur Anbessa Hospital (the country’s largest referral hospital, based in Addis Ababa) alone reaching approximately 100 units per day, a level that is rarely met. O-negative blood was also reported as chronically in short supply. Respondents stated that 56 blood banks were operational nationally at the time of the interview, with an expansion target of 60 by the end of EC 2018 (2025/2026), and that more than 741 hospitals nationwide, including over 150 in Addis Ababa, depended on the national blood supply system.

***<u>Absence of national PBM governance</u>***: all participants reported the lack of a dedicated national PBM policy, guideline, or program. Existing references to PBM were limited to a single chapter in the Appropriate Clinical Use of Blood guideline and a brief mention in the National Surgical, Obstetric, and Anaesthesia Plan. Participants reported relying on institutional guidance, textbooks, or international sources.

***<u>Blood supply constraints:</u>*** All interviewees reported persistent national shortages in the blood supply. The national collection consistently remained below 50% of the estimated annual demand. Component separation was reported to be limited and geographically concentrated, with platelet and O-negative blood widely described as unavailable in routine practice.

To contextualise these constraints at the service level, respondents from the national blood service in Addis Ababa provided operational data. During the reporting period, 61,932 units of whole blood were requested, of which 54,040 units (87.3%) were issued. For red cell components, 43,865 units of concentrated red cells (CRC) were requested, of which 34,945 units (79.7%) were issued. Platelet demand exceeded supply: 38,464 units requested and 31,806 units issued (82.7%), whereas 23,512 units of fresh-frozen plasma were requested and issued in full (100%).

During interviews conducted in November 2025, a Ministry of Health leader reported that national DHIS2 data from the preceding 12 months showed that 145 to 375 major elective surgical procedures were cancelled or referred each month due to blood unavailability. Stakeholders referenced DHIS2 cancellation figures, which aligned with the independently analysed DHIS2 data extracted by the study team.

***<u>Preoperative anaemia management</u>***: reported haemoglobin thresholds for postponing surgery or requiring transfusion ranged from <7 g/dL to <13 g/dL. Intravenous iron and erythropoietin were reported as unavailable. Oral iron was reported as used when time permitted.

***<u>Blood conservation practices:</u>*** Tranexamic acid use was reported by approximately 70% of participants, most frequently in obstetrics/gynaecology and cardiothoracic surgery. Autologous transfusion systems, including cell salvage and acute normovolaemic haemodilution, were not reported as available.

***<u>Blood loss estimation:</u>*** all clinicians reported estimating blood loss using visual assessment, gauze counting, and suction canister measurements. Participants reported that these methods underestimated blood loss by up to 50%.

***<u>Transfusion practice patterns</u>***: participants reported whole blood as the most used product. Blood components were reported to be in limited supply. Over-ordering (wastage) and limited post-transfusion documentation were reported.

***<u>Leadership perspectives:</u>*** all Ministry and hospital leaders reported willingness to adopt PBM if national guidance were issued.

### Triangulated findings across data sources

Facility-level assessments, registry data, and stakeholder interviews showed concordant findings in several domains.

Hospitals reported oxygen availability and access to basic blood products in all facilities, and registry data showed a low perioperative transfusion rate (2.4%).

Laboratory capacity gaps were observed across data sources: facility assessments documented the absence of viscoelastic testing in all hospitals and limited point-of-care testing, while clinicians reported reliance on visual estimation and basic haemoglobin results alone for transfusion decisions.

Medication availability findings were consistent across data sources. Intravenous iron was unavailable at any facility, and clinicians reported relying on oral iron and transfusions to manage anaemia. Tranexamic acid was available in clinical areas and was reported as routinely used by approximately 70% of interviewees.

Blood inventory constraints were reported across all data sources. Facility assessments documented the routine availability of whole blood and packed red blood cells at most sites, whereas clinicians reported national supply constraints and component availability below 15%. Registry data showed that packed red cells were the most frequently used product (61.5%), followed by whole blood (30.8%).

Governance findings were aligned across data sources. Facility assessments identified institutional transfusion policies in all hospitals, while all interviewees confirmed the absence of a national PBM guideline and reliance on institutional or international guidance.

## Discussion

This study provides the first integrated, pre-implementation baseline assessment of the health system for perioperative Patient Blood Management (PBM) in Ethiopia, combining facility level capability, clinical practice, and stakeholder perspectives within a single analytic framework. The findings depict a health system where functional but constrained transfusion services operate in the absence of PBM. Anaemia is largely undiagnosed and untreated preoperatively; blood conservation strategies and haemostatic monitoring are not in place; and transfusion remains a reactive response to blood loss, rather than one component of a coordinated, patient-centred strategy to optimise patients before, during, and after surgery.

Facility assessments revealed strong technical capacity for basic transfusion, including blood typing, emergency access to red cells, and availability of whole blood. However, essential components of PBM were underdeveloped: systematic anaemia detection, access to intravenous iron, point-of-care diagnostics, and blood-loss monitoring technology were frequently unavailable, and basic tests, such as haematological markers (e.g., ferritin) for diagnosing iron deficiency, were often lacking. Advanced haemostatic products, cell salvage, and viscoelastic testing were absent across all facilities. These gaps mirror patterns observed in other low- and middle-income countries, where transfusion services exist but PBM is not yet codified in national policy or financed as a strategic safety intervention.

Importantly, the composite PBM FAT score of 6.6/10 should not be interpreted as evidence of meaningful PBM capability. When disaggregated, the higher scoring domains, blood products, infrastructure, and policy/protocols, reflect general perioperative and transfusion service functionality rather than PBM specific readiness. In contrast, PBM critical domains, systematic anaemia management, intravenous iron availability, haemostatic diagnostics, and structured PBM governance were markedly deficient or absent across all participating facilities. This pattern underscores the need for domain level interpretation rather than reliance on the composite score, particularly in LMIC settings where general perioperative capacity may mask PBM specific gaps.

These findings align with national and regional evidence showing uneven readiness across diagnostic and therapeutic services. Ethiopia’s 2018 service readiness survey reported low availability of hospital transfusion protocols, limited laboratory capacity and weak staff training outside tertiary centres (9). Similar patterns are reported across LMICs, where hospitals frequently depend on locally organised blood supply, experience limited component availability, and report persistent inventory shortages and delays in transfusion (16). When interpreted against international PBM implementation frameworks, Ethiopia remains in an early implementation phase, characterised by technical capacity without national governance or coordinated clinical integration (17).

Placing these findings in a national and continental context strengthens their significance. The Ethiopian Surgical Outcomes Study reported postoperative complications in nearly one in five surgical patients nationwide, despite a young and predominantly low-risk population (8). At the continental level, the African Surgical Outcomes Study (ASOS) showed that African patients experience almost double the global postoperative mortality, with most deaths occurring after surgery (18). Together, these data indicate that excess mortality is driven less by patient risk and more by perioperative system-level gaps. While neither study examined PBM directly, the deficiencies identified in this assessment, limited anaemia management, diagnostic constraints and weak utilisation governance, may be plausible contributors to the failure-to-rescue pathway.

The cohort-level findings from this study require caution. The observed prevalence of preoperative anaemia (7.1%) and transfusion rate (2.4%) are far lower than reported in Ethiopian cohorts, where anaemia prevalence commonly exceeds 40% (19), and 20-40% among reproductive aged women in national and regional estimates (20). The observed perioperative transfusion rate (2.4%) was substantially lower than the 22.7% anticipated from prior literature used to inform the sample size calculation, and should not be interpreted as evidence of appropriate clinical decision-making or effective PBM. In a system characterised by chronic blood shortages, with national collection meeting less than 50% of estimated annual demand, and documented monthly surgical cancellations due to blood unavailability, this figure more plausibly reflects supply-side rationing than patient-centred practice(14). This discrepancy reflects a detection failure: haemoglobin was not systematically measured, and non-random under-ascertainment, whereby testing was most likely in patients already perceived to be at risk, means the reported 7.1% is a documented minimum rather than an epidemiological estimate. External evidence underscores the magnitude of this gap: the African Surgical Outcomes Study (18,20), reported perioperative anaemia in 37.2% of African surgical patients, and Ethiopian-specific studies consistently report preoperative anaemia exceeding 40% in comparable populations (19,20). The registry finding therefore reflects a failure of detection and documentation, not a low burden of disease. Qualitative interviews reinforce this: participants consistently reported the absence of intravenous iron, reliance on oral supplementation when surgery could be delayed, and transfusion as the primary response to anaemia when blood was accessible. The variation in haemoglobin thresholds and reliance on transfusion indicate that anaemia management depends on clinicians rather than protocols, supporting the view that the low reported prevalence of anaemia is due to under-detection rather than effective control.

Inefficient blood utilisation further highlights systemic weakness. Ethiopian studies consistently demonstrate cross-match-to-transfusion ratios well above accepted thresholds, indicating routine over-ordering and suggesting weak coordination between surgical services and blood banks (14,21,22). Notably, these inefficiencies coexist with extreme scarcity. The reported national figures indicated that annual blood collection (∼423,000 units) meets less than half of the estimated national demand (>1 million units), leaving more than 50% of the need unmet, while more than 20% of the national blood demand was reported to be concentrated in Addis Ababa alone. Consistent with these constraints, the Ministry of Health DHIS2, which was independently extracted and analysed by the study team and also referenced by interviewees, indicated that nationally, between 145 and 375 major elective surgical procedures were cancelled or referred each month due to blood unavailability, underscoring how constrained and poorly coordinated blood systems translate into lost surgical access and delayed care, rather than isolated transfusion inefficiency. As with all routinely reported national datasets, DHIS2 figures may be subject to documentation or completeness limitations, but they provide valuable context for understanding system level constraints.

These patterns are more plausibly explained not by clinical need but by the absence of formal blood-ordering schedules, routine audit, and utilisation feedback, as consistently reported by clinicians and blood bank stakeholders in this study and in other settings (10,17). International guidance identifies exactly these behaviours as addressable through blood utilisation governance tools, including maximum surgical blood-ordering schedules (MSBOS), order auditing, and institutional performance monitoring, which, while distinct from PBM’s patient-centred pillars, are complementary system-level stewardship measures relevant to settings of chronic scarcity (10,23). Ethiopia’s current model, which delegates stewardship to individual clinicians within a resource-constrained system, is inherently fragile. Interpretation of the regression findings also requires caution, as the relatively small number of transfusion events limited the precision and stability of multivariable estimates, as reflected by the wide confidence intervals observed for some predictors.

The dominant constraint, however, is institutional rather than technical. The extremely limited number of transfusion medicine specialists identified across participating hospitals further highlights the limited national workforce capacity available to support PBM governance, utilisation oversight, and transfusion stewardship. Ethiopia exhibits the early-stage pattern described by Hofmann and colleagues: strong local enthusiasm, fragmented leadership, no financing framework and limited national coordination (17). The absence of PBM policy ownership, performance indicators, and funding integration is not simply an implementation gap but a recognised structural barrier to national-scale-up (10,17). Priority research should therefore focus on system interventions: linking surgical registries to blood-bank data, piloting bundled PBM models in high-risk populations, implementing national MSBOS tools, and conducting locally grounded economic analyses. Framed against African surgical and national surgical outcome studies, and evidence of avoidable postoperative deaths, PBM must be regarded as a central quality and safety intervention rather than a transfusion policy add-on.

These findings should be interpreted in light of several limitations. The facility assessment was limited to hospitals in Addis Ababa, which may have led to an overestimation of national PBM readiness. Readiness data relied partly on self-report, introducing potential reporting bias despite verification against records where feasible. Preoperative haemoglobin testing and documentation were inconsistent across facilities, likely leading to under-detection and limiting interpretation of the observed prevalence of anaemia. The cross-sectional design reflects readiness at a single point in time and cannot account for dynamic changes in inventories or staffing. Finally, given the non-random under-ascertainment of preoperative haemoglobin, regression estimates involving anaemia as a predictor should be interpreted with particular caution and are not presented as confirmatory findings. The qualitative component also had minor limitations, including the absence of member checking and limited coder triangulation.

In conclusion, this assessment shows Ethiopia has basic blood transfusion capacity but lacks the tools, therapies, and governance for PBM as a preventive care system. Limited access to preoperative anaemia pathways, lack of intravenous iron, and limited diagnostic options lead to reliance on transfusions as a last resort rather than on early anaemia treatment, indicating system-level issues. Chronic blood shortages mean that expanding supply alone can’t meet demand due to untreated anaemia and blood loss. PBM represents a key evidence-based strategy to optimise patient outcomes by preserving and managing the patient’s own blood, which is the WHO’s endorsed goal of PBM. Feasible first steps consistent with the WHO 2024 LMIC PBM toolkit include systematic preoperative anaemia screening, formulary inclusion of intravenous iron, blood utilisation governance, and a national PBM framework. Reduced allogeneic transfusion exposure will follow as a downstream consequence of effective PBM, not as its primary aim. Given rising surgical and obstetric demand, PBM must be prioritised as a core patient-safety strategy, supported by context-specific implementation research.

## Acknowledgment

We gratefully acknowledge all clinicians, hospital administrators, blood bank personnel and national stakeholders who participated in the qualitative interviews and generously shared their experiences and insights. We thank the Ministry of Health, Ethiopia, and the Ethiopian National Blood Bank Service for their collaboration and support in facilitating access to facilities and key informants. We also acknowledge the National Perioperative Quality Improvement Network (NaPQIN) for coordination, operational support and access to perioperative data systems. We are grateful to the World Federation of Societies of Anaesthesiologists for providing the Patient Blood Management Facility Assessment Tool and associated technical guidance, particularly Sara Portugal, Amal Paonaskar, and Matt Rothero, for their invaluable assistance in assessing and securing all the resources needed for this project. We further thank Global Health Partnership for recognising the critical role of patient blood management in health-system strengthening and for providing financial support for this study. Finally, we extend our appreciation to all participating hospitals for their cooperation and commitment to improving perioperative care in Ethiopia.

## Authors’ contributions

The study concept was developed by FKB and TYA. The study was then designed by FKB, TYA, and PKD. Data analysis and interpretation were carried out by TYA and FKB. The first draft of the manuscript was written by FKB, TYA, PKD, TA, FA, JM and AS. The manuscript was critically revised for important intellectual content by all authors. The final draft of the manuscript was reviewed and approved by FKB. Finally, all authors approved the final version of the manuscript.

## Conflict of interest

The authors declare no conflicts of interest.

## Funding

This study was supported by the World Federation of Societies of Anaesthesiologists (WFSA) in collaboration with the Ethiopian Society of Anaesthesiologists Professional Associations (ESAPA) and Project Network for Perioperative and Critical Care (N4PCc), through a grant provided by Global Health Partnerships (Grant number: GHWP2 LG.02). Global Health Partnerships is funded by the UK Department of Health and Social Care (DHSC). The views expressed in this publication are those of the authors and do not necessarily reflect the views of the funders, the UK Government, or any affiliated institutions.

## Data availability statement

De-identified registry data are available from the corresponding author upon reasonable request.

## Notes

### Competing Interest Statement

The authors have declared no competing interest.

### Author Declarations

The Institutional Review Board of the Armauer Hansen Research Institute gave ethical approval for this work (approval number PO-64-23), covering the Ethiopian surgical outcome studystudy including the PBM-FAT facility readiness assessment and stakeholder interviews. The Institutional Review Board of Debre Berhan University, Asrat Woldeyes Health Sciences Campus gave ethical approval for this work (approval number DBU-095), covering the perioperative registry data.

