## Supplementary file for "Perioperative Patient Blood Management (PBM) in Africa: a multicentre mixed-methods assessment of health system readiness and clinical practice in Ethiopia"

**Supplementary Material**

**Table S1**: PBM-FAT scoring framework

Each item in the PBM-FAT tool was assigned a quantitative score according to standardised rules reflecting the availability, timeliness, or presence of key patient blood management (PBM) resources. The scoring approach aimed to convert categorical responses into numeric values suitable for analysis and comparison across facilities.

| **Response category** | **Scoring principle** | **Assigned score** |
| --- | --- | --- |
| Availability | Always / More than half / Less than half / Never | 3 / 2 / 1 / 0 |
| Binary (Yes/No) | Yes / No | 3 / 0 |
| Time to receive critical lab results | <30 min / 30 min–1 hr. / 1–2 hrs. / 2–4 hrs. / >4 hrs. | 3 / 2 / 1 / 0.5 / 0 |
| Time to obtain RBC (emergency) | <15 min / <1 hr. / 1–5 hrs. / 5–10 hrs. / 10–24 hrs. / >24 hrs. | 3 / 2 / 1 / 0.5 / 0.25 / 0 |
| Reliance on an external blood bank | Not at all / Partially reliant / Fully reliant | 3 / 1 / 0 |
| Policy and guideline presence | Yes (local or national) / No | 3 / 0 |
| Staff or equipment count | ≥ 1 / 0 | 3 / 0 |
| Method of record keeping at this facility | If any/none | 3/0 |
| Information/QI/KPI | If at any frequency/ Never | 3/0 |

**Table S2:** Haemoglobin cutoffs to define anaemia severity in individuals

Preoperative anaemia was defined and graded according to the WHO haemoglobin thresholds by age, sex, and pregnancy status (Guideline on haemoglobin cutoffs to define anaemia in individuals and populations: World Health Organisation, 2024).

| **Population** | **Haemoglobin threshold (g/dL) for anaemia** | **Mild anemia** | **Moderate anemia** | **Severe anemia** |
| --- | --- | --- | --- | --- |
| Children 6-59 months of age | <11 | 10.0-10.9 | 7.0-9.9 | <7.0 |
| Children 5-11 years of age | <11.5 | 11.0-11.4 | 8.0-10.9 | <8.0 |
| Children 12-14 years of age | <12 | 11.0-11.9 | 8.0-10.9 | <8.0 |
| Non-pregnant women (>=15 years) | <12 | 11.0-11.9 | 8.0-10.9 | <8.0 |
| Pregnant women | <11 | 10.0-10.9 | 7.0-9.9 | <7.0 |
| Men (>=15 years) | <13 | 11.0-12.9 | 8.0-10.9 | <8.0 |

### PBM-FAT findings

**Table S3:** Infrastructure

| **Variables** | | **Frequency (%)** |
| --- | --- | --- |
| Number of operating theatres | Range (3-22) | |
| Oxygen availability | Yes | 12(100%) |
| ICU or HDU | Yes | 12(100%) |
| Electricity availability | Always | 11(91.7%) |
|  | More than half the time | 1(8.3%) |
| Grid power interruptions | >5 times | 1(8.3%) |
|  | 2-5 times | 6(50.0%) |
|  | 1 time | 4(33.3%) |
|  | Never | 1(8.3%) |
| Backup power availability | Always | 11(91.7%) |
|  | More than half the time | 1(8.3%) |
| Refrigerator for medications in OR complex | Always | 5 (41.7%) |
|  | More than half the time | 1(8.3%) |
|  | Never | 6 (50.0%) |

**Table S4:** Laboratory Testing

| **Variables** | | **Frequency (%)** |
| --- | --- | --- |
| Blood Count (with at least haemoglobin, hematocrit, WBC, and platelets) | Always | 10(83.3%) |
|  | More than half the time | 2(16.7%) |
| Blood count with MCV and MCH | Always | 11(91.7%) |
|  | More than half the time | 1(8.3%) |
| Blood HCO3 level | More than half the time | 2(16.7%) |
|  | Never | 10(83.3%) |
| Blood Lactate level | Always | 3 (25.0%) |
|  | More than half the time | 1 (8.3%) |
|  | Less than half the time | 1 (8.3%) |
|  | Never | 7 (58.3%) |
| Reticulocyte count | Always | 7(58.3%) |
|  | More than half the time | 3(25.0%) |
|  | Less than half the time | 1(8.3%) |
|  | Never | 1(8.3%) |
| Platelet function tests | Always | 7(58.3%) |
|  | More than half the time | 2(16.7%) |
|  | Never | 3(25.0%) |
| Chemistry panel | Always | 9(75.0%) |
|  | More than half the time | 3(25.0%) |
| Blood HCO3 level | More than half the time | 2 (16.7%) |
|  | Never | 10 (83.3%) |
| Blood Lactate level | Always | 3 (25.0%) |
|  | More than half the time | 1 (8.3%) |
|  | Less than half the time | 1 (8.3%) |
|  | Never | 7 (58.30%) |
| Conventional Coagulation Tests (PT, aPTT, INR) | Always | 2 (16.7%) |
|  | More than half the time | 1 (8.3%) |
|  | Less than half the time | 7 (58.3%) |
|  | Never | 2 (16.7%) |
| Other coagulation tests (e.g. Bleeding Time, TT) | Always | 1(8.3%) |
|  | More than half the time | 1 (8.3%) |
|  | Less than half the time | 4(33.3%) |
|  | Never | 6(50.0%) |
| Fibrinogen level | More than half the time | 1 (8.3%) |
|  | Less than half the time | 1 (8.3%) |
|  | Never | 10 (83.3%) |
| Haematinics assessment (Ferritin / Folate / B12) | Always | 1(8.3%) |
|  | More than half the time | 2 (16.7%) |
|  | Less than half the time | 1 (8.3%) |
|  | Never | 8 (66.7%) |
| Serum Iron levels | Always | 2 (16.7%) |
|  | More than half the time | 2 (16.7%) |
|  | Never | 8 (66.7%) |
| Total Iron Binding Capacity (TIBC) | More than half the time | 3 (25.0%) |
|  | Never | 9 (75.0%) |
| Transferrin Saturation (Tsat) level | More than half the time | 1 (8.3%) |
|  | Never | 11(91.7%) |
| Critical result time | <30 minutes | 5 (41.7%) |
|  | 30 minutes-1 hour | 4(33.3%) |
|  | 2-4 hours | 2(16.7%) |
|  | >4 hours | 1(8.3%) |
| **Near-Patient Testing** | | |
| ABG analysis (with at least pH, PaO2 and PaCO2) | More than half the time | 1(8.3%) |
|  | Less than half the time | 2 (16.7%) |
|  | Never | 9(75.0%) |
| Arterial blood gas analysis (with pH, PaO2, PaCO2, and Lactate and BE calculation) | More than half the time | 1(8.3%) |
|  | Less than half the time | 2 (16.7%) |
|  | Never | 9(75.0%) |
| Point-of-care Hgb measurement | Always | 3 (25.0%) |
|  | Less than half the time | 1(8.3%) |
|  | Never | 8(66.7%) |
| Point-of-care ACT measurement | Less than half the time | 1(8.3%) |
|  | Never | 11(91.7%) |
| Point-of-care International Normalized Ratio (INR) measurement | Less than half the time | 3 (25.0%) |
|  | Never | 9 (75.0%) |
| Viscoelastic Tests | Never | 12(100%) |

**Table S5**: Blood Products

| **Variables** | | **Frequency (%)** |
| --- | --- | --- |
| Performs transfusions | Yes | 12(100%) |
| Has a blood bank | Yes | 12(100%) |
| Blood typing availability | Always | 12(100%) |
| Whole blood availability | Always | 10(83.3%) |
|  | More than half the time | 2(16.7%) |
| Packed RBCs availability | Always | 10(83.3%) |
|  | More than half the time | 2(16.7%) |
| Plasma availability | Always | 11(91.7%) |
|  | More than half the time | 1(8.3%) |
| Platelets availability | Always | 8(66.7%) |
|  | More than half the time | 2(16.7%) |
|  | Less than half the time | 2(16.7%) |
| Cryoprecipitate availability | Always | 2(16.7%) |
|  | More than half the time | 1(8.3%) |
|  | Less than half the time | 3(25.0%) |
|  | Never | 6(50.0%) |
| RBC emergency time | <15 minutes | 7(58.3%) |
|  | <1 hour | 5(41.7%) |
| Blood source | An off-site blood bank | 9(75.0%) |
|  | Voluntary unpaid donors at the facility | 3(25.0%) |
| External blood reliance | Fully reliant | 7(58.3%) |
|  | Partially reliant | 3(25.0%) |
|  | Not at all | 2 (16.7%) |

**Table S6**: Policy and Protocols

| **Variables** | | **Frequency (%)** |
| --- | --- | --- |
| Transfusion policy | Yes, local/facility policy | 6(50.0%) |
|  | Yes, national policy | 6(50.0%) |
| Single unit policy | Yes | 9(75.0%) |
|  | No | 3(25.0%) |
| Major hemorrhage protocol | Yes | 8(66.7%) |
|  | No | 4(33.3%) |
| Transfusion or hemovigilance committee | Yes | 10(83.3%) |
|  | No | 2(16.7%) |
| Monitor transfusion rates | Yes | 12(100%) |
| PBM program | Yes | 6(50.0%) |
|  | No | 6(50.0%) |

**Table S7**: Surgical service and workforce

| **Variables** | | **Frequency (%)** |
| --- | --- | --- |
| Surgical services | General/GI | 9(75.0%) |
|  | Trauma | 7(58.3%) |
|  | Orthopedics | 8(66.7%) |
|  | Obstetrics/Gynaecology | 10(83.3%) |
|  | Neurosurgery | 7(58.3%) |
|  | Plastic/reconstructive | 6(50.0%) |
|  | Urology | 8(66.7%) |
|  | Otolaryngology (ENT) | 7(58.3%) |
|  | Cardiac | 4(33.3%) |
|  | Thoracic | 5(41.7%) |
|  | Vascular | 7(58.3%) |
|  | Pediatric | 8(66.7%) |
|  | Maxillofacial | 7(58.3%) |
|  | Other | 4(33.3%) |
| Major surgery frequency | Daily | 11(91.7%) |
|  | Weekly | 1(8.3) |
| **Service types** | **Total # in all hospitals** | **Ranges** |
| **Anesthesia services** | | |
| Specialist physician anaesthesia providers | 67 | (1-15) |
| Non-specialist physician anaesthesia providers | 170 | (0-42) |
| Nurse anaesthesia providers | 86 | (0-47) |
| Other (non-physician, non-nurse) anaesthesia providers | 92 | (0-32) |
| **Surgical services** | | |
| Physician surgeons | 408 | (0-142) |
| General Doctors who provide surgery (excluding obstetrics) | 68 | (0-68) |
| Non-physicians who provide surgery (excluding obstetrics) | 2 | (0-2) |
| **Obstetrics/Gynaecology Services** | | |
| Obs/gyn physicians | 152 | (0-48) |
| GP who provides CS | 0 | NA |
| Non-physicians who provide CS | 4 | (0-2) |
| Midwives | 677 | (0-145) |
| **Other services** | | |
| Critical Care physicians | 108 | (0-54) |
| Theatre (scrub) nurses | 501 | (4-123) |
| Hematologists | 24 | (0-8) |
| Transfusion medicine physicians | 4 | (0-4) |
| Laboratory technicians | 392 | (3-70) |
| Pediatricians | 144 | (0-42) |
| Pharmacists | 630 | (19-95) |

**Table S8**: Medications

| **Variables** | | **Frequency (%)** |
| --- | --- | --- |
| IV Tranexamic acid (TXA) | Always | 8(66.7%) |
|  | More than half the time | 2(16.7%) |
|  | Never | 2(16.7%) |
| Other antifibrinolytics | Always | 2(16.7%) |
|  | Never | 10(83.3%) |
| IV Iron | Always | 3(25.0%) |
|  | Never | 9(75.0%) |
| IV Vitamin B complex | Always | 12(100%) |
| Vasopressors | Always | 5(41.7%) |
|  | More than half the time | 2(16.7%) |
|  | Less than half the time | 4(33.3%) |
|  | Never | 1(8.3%) |
| Inotropes | Always | 10(83.3%) |
|  | Less than half the time | 2(16.7%) |
| Heparins | Always | 11(91.7%) |
|  | More than half the time | 1(8.3%) |
| Protamine | Always | 1(8.3%) |
|  | More than half the time | 1(8.3%) |
|  | Less than half the time | 1(8.3%) |
|  | Never | 9(75.0%) |
| Recombinant Factor VIIa | Always | 1(8.3%) |
|  | Less than half the time | 2(16.7%) |
|  | Never | 9(75.0%) |
| Prothrombin Complex Concentrate (PCC) | Less than half the time | 2(16.7%) |
|  | Never | 10(83.3%) |
| Fibrinogen Concentrate (FC) | Less than half the time | 2(16.7%) |
|  | Never | 10(83.3%) |
| Activated charcoal | Less than half the time | 3(25.0%) |
|  | Never | 9(75.0%) |
| Desmopressin (DDAVP) | Less than half the time | 2(16.7%) |
|  | Never | 10(83.3%) |
| IV Vitamin K | Always | 10(83.3%) |
|  | Never | 2(16.7%) |
| IM Vitamin K (for neonates) | Always | 9(75.0%) |
|  | More than half the time | 1(8.3%) |
|  | Never | 2(16.7%) |
| IV Calcium Gluconate | Always | 10(83.3%) |
|  | More than half the time | 2(16.7%) |
| Topical haemostatic agents | Less than half the time | 2(16.7%) |
|  | Never | 10(83.3%) |
| Oral Iron | Always | 10(83.3%) |
|  | More than half the time | 2(16.7%) |
| Oral Folic Acid supplements | Always | 12(100%) |
| Oral Vitamin B12 supplements | Always | 7(58.3%) |
|  | Less than half the time | 3(25.0%) |
|  | Never | 2(16.7%) |
| Oral Vitamin D supplements | Always | 5(41.7%) |
|  | More than half the time | 2(16.7%) |
|  | Less than half the time | 1(8.3%) |
|  | Never | 4(33.3%) |
| Oral Vitamin K supplements | Always | 1(8.3%) |
|  | Less than half the time | 2(16.7%) |
|  | Never | 9(75.0%) |
| Erythropoiesis-stimulating agents (ESAs) | Always | 3(25.0%) |
|  | Never | 9(75.0%) |
| Antiplatelet agents | Always | 11(91.7%) |
|  | More than half the time | 1(8.3%) |

**Table S9**: Equipment

| **Variables** | | **Frequency (%)** |
| --- | --- | --- |
| Basic monitoring devices | Always | 7(58.3%) |
|  | More than half the time | 5(41.70%) |
| End-tidal CO2 monitoring | Always | 2(16.7%) |
|  | More than half the time | 3(25.0%) |
|  | Less than half the time | 3(25.0%) |
|  | Never | 4(33.3%) |
| Intra-arterial BP monitoring equipment | Always | 1(8.3%) |
|  | More than half the time | 1(8.3%) |
|  | Less than half the time | 2(16.7%) |
|  | Never | 8(66.7%) |
| Infusion or syringe pump devices | Always | 3(25.0%) |
|  | Less than half the time | 4(33.3%) |
|  | Never | 5(41.7%) |
| Rapid infusion devices | Always | 3(25.0%) |
|  | Less than half the time | 3(25.0%) |
|  | Never | 6(50.0%) |
| Refrigerator | Always | 5(41.7%) |
|  | More than half the time | 1(8.3%) |
|  | Never | 6(50.0%) |
| Simple autologous autotransfusion | Never | 12(100%) |
| Low-tech autologous autotransfusion system | Never | 12(100%) |
| High-tech autologous Cell Saver System | Less than half the time | 1(8.3%) |
|  | Never | 11(91.7%) |
| Thermometer for intermittent temperature monitoring | Always | 8(66.7%) |
|  | Less than half the time | 3(25.0%) |
|  | Never | 1(8.3%) |
| Device for continual temperature monitoring | Always | 6(50.0%) |
|  | Less than half the time | 3(25.0%) |
|  | Never | 3(25.0%) |
| Electric warming blanket or device | Always | 3(25.0%) |
|  | Less than half the time | 1(8.3%) |
|  | Never | 8(66.7%) |
| Warming mattress | Less than half the time | 1(8.3%) |
|  | Never | 11(91.7%) |
| Fluid warming device (counter current system) | Always | 2(16.7%) |
|  | Less than half the time | 1(8.3%) |
|  | Never | 9(75.0%) |
| Surgical suction | Always | 8(66.7%) |
|  | More than half the time | 3(25.0%) |
|  | Never | 1(8.3%) |
| Surgical Tourniquets | Always | 5(41.7%) |
|  | Less than half the time | 6(50.0%) |
|  | Never | 1(8.3%) |
| Electrocautery (Diathermy) | Always | 9(75.0%) |
|  | More than half the time | 3(25.0%) |
| Biomedical services availability | Always | 12(100%) |

**Table S10**: Perioperative Services

| **Variables** | | **Frequency (%)** |
| --- | --- | --- |
| Spinal anesthesia availability | Always | 12(100%) |
| General anesthesia availability | Always | 12(100%) |
| Patient evaluation by anesthesia | Always | 12(100%) |
| Pre-operative assessment for elective cases | Always | 11(91.7%) |
|  | Never | 1(8.3%) |
| Anemia clinic | Yes | 1(8.3%) |
|  | No | 11(91.7%) |
| IV Iron availability | Not available | 12(100%) |
| WHO Surgical Safety Checklist | Always | 12(100%) |
| Bleeding risk assessed and/or discussion | Always | 7(58.3%) |
|  | More than half the time | 2(16.7%) |
|  | Less than half the time | 2(16.7%) |
|  | Never | 1(8.3%) |
| Interventional radiology/radiological embolisation availability | Not available | 12(100%) |

**Table S11**: Surgical Volume

|  | **Total # in all hospitals** | **Ranges** |
| --- | --- | --- |
| Total_surgical_cases_12mo | 83,085 | (0-19553) |
| Elective_cases_12mo | 44,597 | (0-14329) |
| Csection_cases_12mo | 27,998 | (0-5189) |
| Polytrauma_cases_12mo | 2408 | (0-1121) |
| Note: some of the hospitals have no data on the number of electives, and polytrauma. In addition, some hospitals only provide CS services. | | |

**Table S12**: Information Management, quality and safety

| **Variables** | | **Frequency (%)** |
| --- | --- | --- |
| Record-keeping method | Both | 5(41.7%) |
|  | Electronic | 7(58.3%) |
| QI meetings frequency | Daily/Weekly | 2(16.7%) |
|  | Monthly | 10(83.3%) |
| Surgical deaths tracking | Daily/Weekly | 5(41.7%) |
|  | Monthly | 4(33.3%) |
|  | As needed | 2(16.7%) |
|  | Never | 1(8.3%) |
| Surgical morbidity & mortality report | Monthly | 11(91.7%) |
|  | As needed | 1(8.3%) |
| Data prospectively collected for patient outcomes | Daily/Weekly | 3(25.0%) |
|  | Monthly | 7(58.3%) |
|  | As needed | 1(8.3%) |
|  | Never | 1(8.3%) |
| KPI tracking | Daily/Weekly | 1(8.3%) |
|  | Monthly | 11(91.7%) |
